# Changes in socioeconomic differences in drinking patterns before and after retirement - a 22-year follow-up study

**DOI:** 10.1101/2025.09.28.25336308

**Authors:** Karjala Anni, Salonsalmi Aino, Lahelma Eero, Rahkonen Ossi, Lallukka Tea

## Abstract

**Background:** Population is rapidly ageing globally, and alcohol consumption among older adults has increased in recent years. Alcohol consumption is known to be socioeconomically patterned, but little is known about how these socioeconomic differences evolve as individuals age and over the transition into statutory retirement.

**Objective:** We aimed to examine changes in socioeconomic differences in drinking patterns during a 22-year follow-up among retiring employees.

**Methods:** We analyzed changes in drinking patterns among a cohort of ageing employees using data from the Helsinki Health Study surveying the aging City of Helsinki employees. Data were collected in five phases between 2000 and 2022. All participants were 40-60-year-old in Phase 1 and retired during the follow-up. The total number of participants was 4889. Non-drinking, binge drinking and average weekly units were used as measures for alcohol consumption. We used occupational class as our indicator of socioeconomic position and evaluated changes in alcohol consumption using generalized linear mixed effect models (GLMM). Age and gender were added as covariates.

**Results:** The higher class drunk more but the lower class were more likely to be binge drinkers or non-drinkers. During ageing, alcohol consumption by weekly doses and binge drinking decreased, and non-drinking became more common in all socioeconomic groups. The socioeconomic differences persisted over the follow-up, with little change over retirement transition.

**Conclusions:** Alcohol drinking declined, and non-drinking became more common, with socioeconomic differences in midlife persisting in older adulthood and over retirement transition.

## Introduction

The Western world is entering an unprecedented demographic shift, with the number of people aged 65 and older set to double by mid-century^1^. Alcohol use is a major global risk factor for injuries, numerous non-communicable diseases, and premature mortality^2^. In Western countries, consumption has been rising particularly among older adults in recent years due to older cohorts drinking more than younger ones ^3–5^. This trend is concerning, as ageing groups are especially vulnerable to the harmful effects of alcohol^5^. Contradictory to previous perception, no safe levels for alcohol use are known^6^, as even small doses may increase the risks for various health conditions, including stroke and cancer^7,8^.

There is a well-established socioeconomic gradient in alcohol drinking^9,10^. Alcohol drinking accounts for a considerable part of socioeconomic health inequalities^11^, and narrowing socioeconomic differences in alcohol use have started increasing again in Finland in recent years^12^. Although individuals in low socioeconomic positions are more often non-drinkers and consume less alcohol compared to individuals in high socioeconomic positions^13^, they experience greater alcohol-related harm^9,13,14^. This discrepancy between alcohol consumption and alcohol-related harm is called the alcohol harm paradox^14^.

Alcohol use changes during life-course, typically reaching a peak in early adulthood, then declining toward older adulthood^15^. Therefore, socioeconomic differences in drinking patterns might also change with ageing. Existing evidence on the topic has largely focused on trajectory models, where latent drinking patterns and associations of socioeconomic indicators with these trajectories have been examined. Overall, according to these studies, higher education and income are consistently linked with more frequent or increasing alcohol use in midlife and older age^16–19^, but no studies have looked at how socioeconomic differences in drinking patterns develop during ageing.

Retirement is a key life transition that may influence alcohol use during ageing. A recent review concluded that the overall impact of retirement on alcohol consumption remains unclear^20,^ with studies reporting increases^21,22^, decreases^19,23^ or no change^17,24^. In the French GAZEL cohort, statutory retirement was followed by a temporary rise in heavy drinking, which persisted for female managers throughout the 5-year follow-up^25^. On the other hand, a Finnish public sector study found that while most individuals maintained stable risky drinking patterns after retirement, a subgroup of particularly those in lower socioeconomic positions temporarily increased their alcohol use with retirement, whereas those from higher socioeconomic groups were more likely to belong to the slowly decreasing risky drinking group with overall higher risky drinking levels ^27.^ A Swedish study found that those who retired increased their weekly alcohol consumption, and that this increase was driven by those with higher education^28^.

Although trajectories of alcohol use in older adulthood have been investigated, there are no previous studies looking at how differences in alcohol use develop by socioeconomic group during ageing and over the retirement transition. To address this gap in literature, we examined changes in socioeconomic differences in drinking patterns during a 22-year follow-up and over retirement transition among ageing and retiring public sector employees.

## Materials and methods

### Study population

This study is part of the Helsinki Health Study (HHS), a longitudinal survey-based cohort of the employees of the City of Helsinki, Finland. The HHS investigates health and well-being of the participants from adulthood and midlife to older age^29.^ Phase 1 survey was conducted between 2000 and 2002 and targeted all employees of the City of Helsinki in Finland who turned 40, 45, 50, 55, or 60 during that period. Of these, 8,960 responded (response rate: 67%). Approximately 80% of respondents were women, reflecting the gender distribution among the employees of the City of Helsinki^29^. Follow-up surveys were conducted in 2007 (Phase 2: 83%, N = 7332), 2012 (Phase 3: 79%, N = 6814), 2017 (Phase 4: 82%, N = 6832), and 2022 (Phase 5: 75%, N = 5944). Data collection was carried out via mailed questionnaires, with up to three reminder letters sent to non-respondents. Non-response analyses have been conducted to confirm the representativeness of the data for the target population^29,30^.

Those participants who retired statutorily during the follow-up and had provided information on the time of retirement were considered (n=5512). Those who were absent from work due to other reasons or retiring due to disability were not included. A small number of participants who lacked data at all phases for weekly doses (n=103), binge drinking (n=35) and non-drinking (n=16) items were excluded from each analyses, but participants who had responded at each item at least once during the follow-up were included in the analysis, thus resulting in a different number of participants in analyses for each drinking habit. The final number of participants considered for the analyses was 4889.

### Alcohol measures

Average weekly alcohol consumption of alcohol was asked by three questions, each having seven response alternatives. The first question inquired after the average number of bottles of beer and cider per week, the second one the average number of bottles of wine or other mild beverages per week and the third one the average the number of bottles of spirits per month. From these, the average consumption of alcohol units per week was calculated (where a unit of alcohol was defined as 12 grams of pure alcohol).

Binge drinking was defined as consuming six or more units of alcohol on a single occasion. The questionnaires included a question with six possible responses, asking how often the respondent drank at least this amount. Those who drank six or more alcohol units on a single occasion once per month or more often were considered as binge drinkers.

Non-drinking was derived from a question regarding the current frequency of drinking, which was asked with ten response alternatives. By combining the alternatives, respondents were divided into five classes. The classes were non-drinkers, those drinking a few times per year, those drinking a few times per month, those drinking a few times per week and those drinking more often. The non-drinkers class also included those drinking once a year or less often. Our non-drinking variable included ex-drinking.

### Retirement

In modeling retirement, the x-axis was modified to present the time difference between the age at the particular phase and the actual retirement age of the participant with retirement time as the reference point (time zero), allowing for the distinction between the years before and after retirement. Thus, the time variable was from –22 to 22 before and after retirement. However, due to few participants at the very extremes, we included time from –20 to 20. Furthermore, we analyzed pre-and post-retirement slopes for different SEP groups.

### Occupational class

We divided the participants into three occupational classes, following previous procedures^31^. Occupational class at phase 1 was used as a determinant for the socioeconomic position. The highest class “professionals” included managers and professionals, occupations that required university-level qualifications or were classified as managerial positions and involve mainly autonomous managerial or supervisory tasks (titles such as doctors or teachers). Those occupations requiring college-level qualifications and involving both supervisory and routine tasks with less autonomy were included in the “semi-professionals” (titles such as nurses, foremen, and technicians).

The third occupational class “routine and manual workers” consisted of routine non-manual employees, requiring vocational training or no specific qualifications, and involving non-supervisory clerical and other non-manual tasks (titles such as child minders and health care assistants) and manual workers, also requiring vocational training or no specific qualifications (those working for example in transportation or cleaning).

### Covariates

Baseline age and gender were used as covariates in the study. Data on self-rated health were obtained from the Phase 1 survey. Age and gender (women/man) were obtained from the questionnaire.

### Statistical analyses

To present the descriptive characteristics of the participants, data were presented as means, medians or as counts with percentages. We used generalized linear mixed models (GLMMs) to analyze changes in average weekly portions by occupational classes, as this data was non-normally distributed with a right skew. Since the gamma model with a log-link function does not accommodate zero values, values of zero were reassigned a value of 0.1. In the unadjusted models, alcohol consumption over study phases were used as the dependent variable and added the occupational class and an interaction between time and occupational class as a fixed effect in the models. Second, to investigate how non-drinking or binge drinking behaved during agreeing, we modeled a binary GLMM with a logit-link function. The predicted probabilities were plotted from these models. A subject-specific intercept was included as a random effect. In the analyses regarding weekly units, random intercept and slope were included. In the analyses regarding binge drinking, only random intercept was included in the models. Adjusted models included age and gender as fixed effects.

Furthermore, to evaluate whether changes in alcohol use patterns differed by socioeconomic position across the retirement transition, we estimated separate slopes for the pre- and post-retirement periods. This was achieved by including two-way interaction terms between socioeconomic indicators and time within each period (i.e., SES x preretirement and SES x postretirement). In this way, we were able to compare the rate of change in each socioeconomic group before and after retirement. Analyses were conducted using SPSS version 29.

## Results

Descriptive results for the study participants at Phase 1 are presented in Table 1.

**Table 1.** Descriptive results of the Helsinki Health Study participants in 2000–2002 (Phase 1), n=4889.

|  | **Professionals**<br>**(n=1611)** | **Semi-professionals**<br>**(n=916)** | **Manual & Routine**<br>**Workers (n=2362)** | **Total**<br>**(n=4889)** | **n** |
| --- | --- | --- | --- | --- | --- |
| **Age, Mean (SD)** | 52.77 (5.23) | 52.11 (5.28) | 52.48 (4.90) | 52.51 (5.08) | 4889 |
| **Women, n (%)** | 1157 (71.8%) | 719 (78.5%) | 2070 (87.6%) | 3946<br>(80.7%) | 4889 |
| **Mean Weekly**<br>**Portions (SD)** | 6.54 (8.28) | 4.98 (6.30) | 3.83 (5.18) | 4.95 (6.68) | 4709 |
| **Non-drinkers (%)** | 106 (6.6%) | 83 (9.1%) | 340 (14.4%) | 523 (10.8%) | 4822 |
| **Binge drinkers (%)** | 351 (21.8%) | 186 (20.3%) | 433 (19.2%) | 970 (20.3%) | 4782 |

For weekly doses, professionals had higher consumption than those in lower socioeconomic groups throughout the follow-up (Figure 1), and these socioeconomic differences were statistically significant (p<0.001). Weekly doses declined over time across all groups, and the overall interaction was not significant (SEP x time, p=0.152), indicating that socioeconomic differences in weekly doses remained stable over the follow-up. Men consumed more weekly doses than women (b=0.779, 95% CI 0.709– 0.850, p<0.001), but the rates of decline in weekly doses was similar across genders. No changes in the trajectory around retirement age (time=0) were observed.

**Figure 1.**
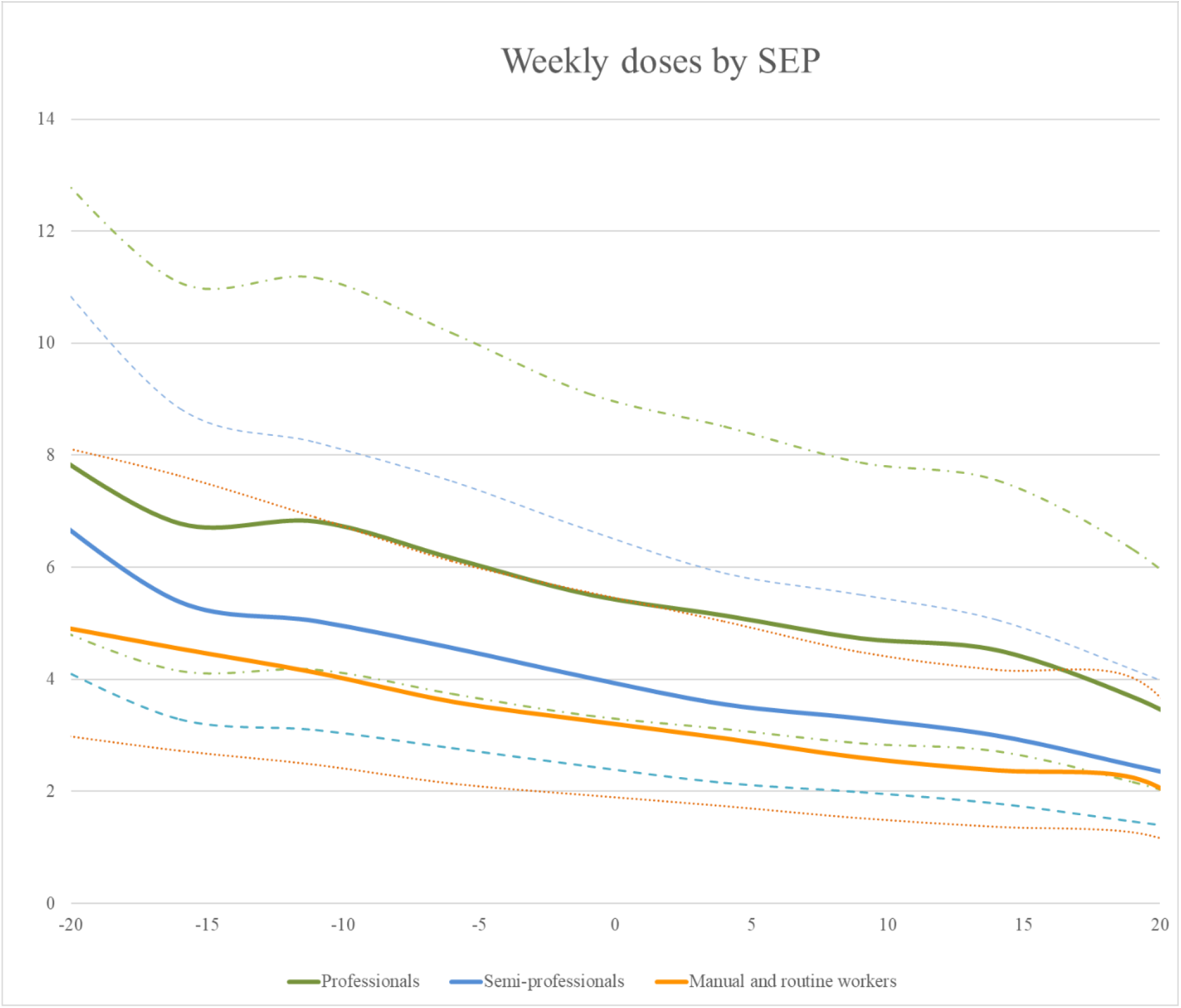
Model predicted weekly doses and 95% CI’s in socioeconomic groups before and after retirement (year 0) (n=4709). Adjusted for age and gender.

**Table 2.** Model coefficients (with 95% CI, log scale) for weekly doses by socioeconomic position (SEP), interactions with time, and covariates among retiring former employees of the Helsinki Health Study (n = 4709)

| Model term | Coefficient (b) | 95% CI (Lower, Upper) | p-value |
| --- | --- | --- | --- |
| **SEP (p=<0.001 for overall interaction at time=0)** |  |  |  |
| Professionals | 0.378 | 0.315, 0.440 | <0.001 |
| Semi-professionals | 0.148 | 0.073, 0.223 | <0.001 |
| Manual and routine workers | ref | ref | ref |
| **Time x SEP (p=0.152 for overall interaction)** |  |  |  |
| Time x Professionals | -0.023 | -0.025, -0.021 | <0.001 |
| Time x Semi-professionals | -0.025 | -0.027, -0.022 | <0.001 |
| Time x manual and routine workers | -0.022 | -0.024, -0.020 | <0.001 |
| **Covariates** |  |  |  |
| Age at baseline | -0.003 | -0.008, 0.003 | 0.312 |
| Gender (ref: women) | 0.779 | 0.709, 0.850 | <0.001 |
| Time x Gender (ref: women) | 0.000 | -0.002, 0.003 | 0.780 |

Binge drinking declined over time across all socioeconomic groups (Figure 2). Overall, manual and routine workers were slightly more likely to be binge-drinkers than the other socioeconomic groups, but the overall SEP x time interaction was not significant (p=0.472), indicating that socioeconomic differences remained similar over follow-up.

**Figure 2.**
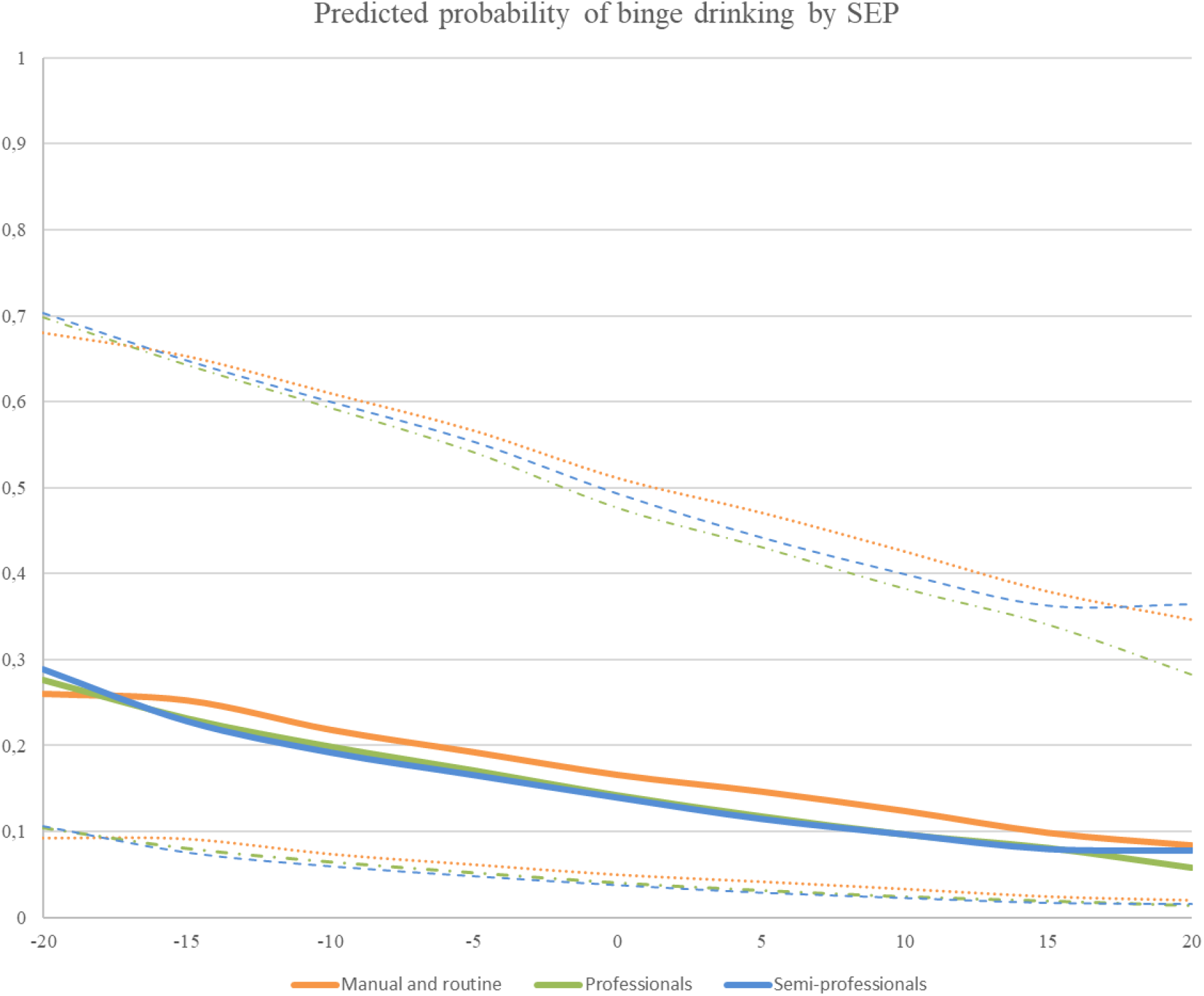
Model predicted probability and 95% CI’s of binge drinking in socioeconomic groups before and after retirement (year 0) (n=4782). Adjusted for age and gender.

Men were much more likely to binge drink than women (b=1.853, 95% CI 1.676 to 2.030, p<0.001). Furthermore, the decline in binge drinking was somewhat faster among men than women (b=-0.012, 95% CI from -0.025 to 0.001), but this interaction was non-significant (p=0.069). Higher baseline age was associated with slightly lower likelihood of binge drinking (b=-0.024, 95% CI from -0.040 to 0.08, p=0.004). Trajectories showed little change around retirement age.

Professionals and semi-professionals were less likely to report non-drinking compared with manual and routine workers (p<0.001) (Figure 3). Non-drinking increased over time in all groups with the interaction of SEP with time being statistically non-significant (p=0.204), suggesting stable socioeconomic differences over follow-up. Men were less likely to be non-drinkers than women (b=-0.289, 95% CI -0.505 to -0.072, p= 0.009), but the time x gender interaction was non-significant (p=0.293) suggesting similar change for men and women. There was no marked change around retirement age in non-drinking.

**Table 3.** Model coefficients (with 95% CI, log scale) for binge drinking by socioeconomic position (SEP), interactions with time, and covariates among retiring employees from the Helsinki Health Study (n = 4782)

| Model term | Coefficient (b) | 95% CI (Lower, Upper) | p-value |
| --- | --- | --- | --- |
| **SEP (overall $p=0.083$ at time=0)** |  |  |  |
| Professionals | -0.131 | -0.305, 0.043 | 0.141 |
| Semi-professionals | -0.226 | -0.438, -0.014 | 0.037 |
| Manual & routine workers | ref | ref | ref |
| **Time x SEP (overall p=0.472)** |  |  |  |
| Professionals x time | -0.048 | -0.060, -0.037 | <0.001 |
| Semi-professionals x time | -0.059 | -0.073, -0.044 | <0.001 |
| Manual & routine workers x time | -0.052 | -0.061, -0.042 | <0.001 |
| **Covariates** |  |  |  |
| Gender (ref: women) | 1.853 | 1.676, 2.030 | <0.001 |
| Time x Gender (ref: women) | -0.012 | -0.025, 0.001 | 0.069 |
| Age | -0.024 | -0.040, -0.008 | 0.004 |

**Figure 3.**
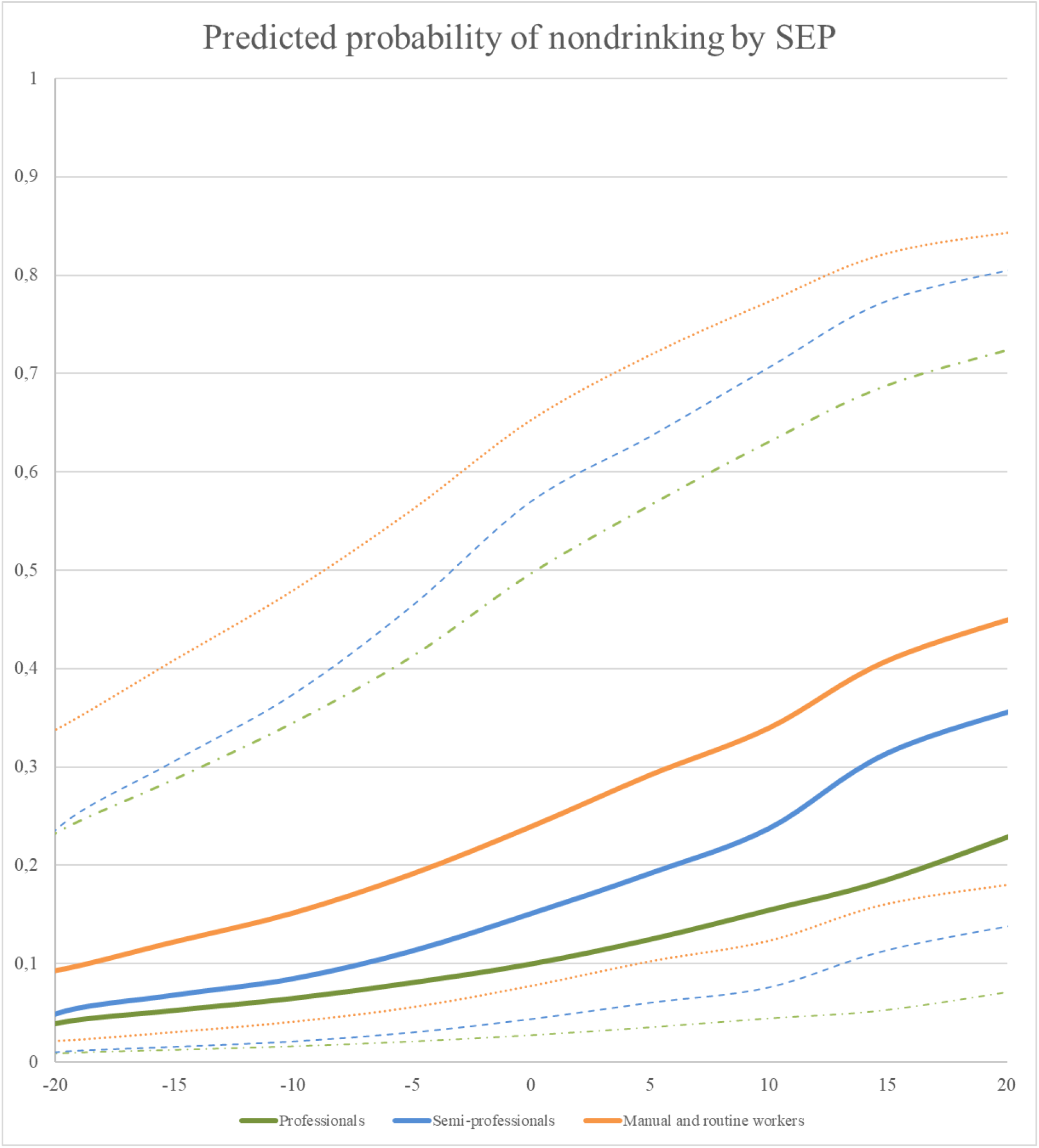
Model predicted probability of non-drinking and 95% CI’s in socioeconomic groups before and after retirement (year 0) (n=4822). Adjusted for age and gender.

**Table 4.** Coefficients (with 95% CI, log scale) for non-drinking by socioeconomic position (SEP), interactions with time, and covariates among retiring employees (n = 4782)

| Model term | Coefficient (b) | 95% CI (Lower, Upper) | p-value |
| --- | --- | --- | --- |
| **SEP (overall p=<0.001 at time=0)** |  |  |  |
| Professionals | -1.195 | -1.386, -1.004 | <0.001 |
| Semi-professionals | -0.742 | -0.959, -0.525 | <0.001 |
| Manual & routine workers | ref | ref | ref |
| **Time x SEP overall (p=0.204)** |  |  |  |
| Professionals x time | 0.086 | 0.072, 0.099 | <0.001 |
| Semi-professionals x time | 0.103 | 0.088, 0.119 | <0.001 |
| Manual & routine workers x time | 0.095 | 0.086, 0.103 | <0.001 |
| **Covariates** |  |  |  |
| Gender (ref: women) | -0.289 | -0.505, -0.072 | 0.009 |
| Time x Gender (ref: women) | -0.009 | -0.026, 0.008 | 0.293 |
| Age at baseline | 0.022 | 0.039, 0.006 | 0.009 |

Additional analyses of retirement-related changes in alcohol use indicated little evidence of systematic change with retirement, with period x time interaction being nonsignificant for all drinking patterns across socioeconomic groups (supplementary table 1). For weekly doses, slopes were close to zero and nonsignificant in all occupational classes both before and after retirement, suggesting stable levels of consumption across the transition. Similarly, non-drinking rates remained essentially unchanged with retirement, with all slopes small and nonsignificant. The only notable change was a significant pre-retirement decline in binge drinking among professionals (–0.013, 95% CI –0.22 to – 0.03, p = 0.009), which did not continue after retirement. In other groups, pre- and post-retirement changes were minor and nonsignificant. Overall, these findings indicate that alcohol use trajectories remained largely stable across the retirement transition, with no consistent evidence of either increases or decreases by occupational class.

## Discussion

Alcohol use among ageing participants declined over the 22 follow-up years. We found that occupational class differences in alcohol use remained largely stable over the follow-up. Participants in higher socioeconomic positions consumed more weekly doses and were less often binge drinkers or non-drinkers as compared to those in lower socioeconomic position. Across all groups, consumption as weekly units and binge drinking declined and non-drinking increased with age, with socioeconomic differences remaining similar throughout follow-up. To our knowledge, this is the first study to examine changes in alcohol use patterns across socioeconomic groups in an ageing and retiring occupational cohort. The findings add to existing evidence that socioeconomic position not only shapes overall levels of drinking but that the differences observed in mid-life persist into older adulthood.

The findings regarding overall patterns in alcohol use observed in this study are in line with previous evidence suggesting a decrease in units and binge drinking and increase in non-drinking during ageing^15,16^. For persisting socioeconomic differences for weekly units, previous evidence investigating latent trajectories of alcohol use have found that higher education and income consistently associate with stable or increasing alcohol use in midlife and older age^16–19^. Although these studies did not directly examine how socioeconomic differences develop, their findings can be interpreted as parallel to our findings of persistingly higher weekly doses in higher socioeconomic groups, although no changes in socioeconomic differences were observed^17^.

Binge drinking was more common in the manual and routine workers group, which is in line with previous reports that have found those in lower socioeconomic groups being binge drinkers more often^10,32^. However, there is also previous evidence indicating that older women in higher socioeconomic positions may be especially prone to heavier episodic drinking, which may explain that the observed SEP differences were small, as our cohort mainly consists of ageing women^32,33^ . Binge drinking decreased in all groups over follow-up, which is in line with previous findings reporting decreases in binge drinking with age^16^. Nondrinking was more common in manual and routine workers and increased in all groups over follow-up. The overall pattern is in line with previous evidence, with those in lower socioeconomic positions being non-drinkers more often^13^, and non-drinking increasing during ageing^16^.

We found no marked changes in socioeconomic differences in drinking patterns around retirement age. These patterns are broadly in line with previous research reporting stability or only minor or temporary changes in alcohol consumption around retirement^17,27^. They are, however, in contrast with findings from Finnish and French studies, which found changes in risky or heavy drinking retirement especially in lower socioeconomic groups^25,28^ and a Swedish study which found that those who retired increased their weekly alcohol consumption, and that this increase was driven by those with higher education^28^. The difference may stem from slightly different alcohol measures, or cultural differences. Nevertheless, the results concerning changes in alcohol use with retirement transition warrant further research.

Overall, the persistence of socioeconomic differences in alcohol use over follow-up may reflect a combination of different mechanisms. First, drinking behaviors in adulthood are often characterized by habitual stability. Second, the reasons underlying socioeconomic differences might change during ageing, and contribute to alcohol use in socioeconomic groups universally. These factors could be things such as declining biological tolerance to alcohol, medication use and greater health awareness in later life. Finally, selective attrition may also play a role. Heavier drinkers in lower socioeconomic positions were more likely to drop out, which can leave a healthier and lighter-drinking subset in follow-up, dampening possible changes in socioeconomic differences

It is worth noting that non-drinking likely reflects different mechanisms across socioeconomic groups. Our non-drinking variable included ex-drinking. In lower SEP, non-drinking may include more former drinkers and ill-health quitters, and thus non-drinking may result from structural disadvantage and health selection^34,35^. By contrast, in higher SEP groups nondrinking may more often be a deliberate, health-conscious choice, also reflecting social norms that value moderation^36^. This heterogeneity should be taken into account when interpreting the results; non-drinking may indicate different backgrounds and situations.

## Strengths and limitations

A key strength of this study is the use of a large, occupational cohort with repeated measurements over an extended follow-up and a good response rate, allowing us to capture changes across midlife and older adulthood and over retirement transition. This longitudinal design enabled us to take into account both group-level trends and individual variation in drinking behavior over time. Another strength is the simultaneous consideration of multiple alcohol indicators, which describe different dimensions of alcohol use, known to vary by socioeconomic groups.

However, some limitations must be acknowledged. Alcohol use was - as it always is - self-reported, which may be prone to underreporting, particularly among heavy drinkers. Furthermore, social approval bias may also play a role, with individuals tending to overreport behaviors perceived as healthy and underreport those viewed as unhealthy^37^. Moreover, biases in reporting may not be randomly distributed: socioeconomic differences in knowledge, health literacy, or cultural norms could lead to systematic variation in reporting accuracy across groups, which in turn may influence the observed socioeconomic gradients in health behaviors. The cohort of Helsinki municipal employees reflects the socioeconomic and gender structure of the Finnish public sector, but findings may not fully generalize to the general population. The predominance of women (80%) may also affect the transferability of results, as women’s drinking patterns and socioeconomic gradients in alcohol use differ from those of men. Nevertheless, the study population provides valuable insights into an ageing workforce within a Nordic welfare state context, where alcohol is a key driver of health inequalities.

Although response rates in this cohort were relatively high, missing data accumulated across survey waves. GLMM is robust to data missing at random and enabled us to use all available information, reducing the likelihood of attrition bias. However, selective dropout was observed for binge drinking. To address this, we compared baseline characteristics of responders and non-responders across follow-ups. Attrition was patterned by both SEP and binge drinking, with lower SEP and binge drinkers dropped out more often. For other drinking habits, no such differences were observed. While the missingness pattern is broadly consistent with missing at random (MAR), but selective dropout of heavier drinkers in lower SEP groups raises the possibility of missing not at random (MNAR). Consequently, the observed trajectories of binge drinking over follow-up may underestimate true levels, especially in lower SEP groups and in those with heavier alcohol use.

It is also important to consider potential age-period-cohort effects when interpreting these findings. Declines in alcohol consumption with ageing are well documented, reflecting factors such as physiological changes, increasing health concerns, and medical contraindications in later life. Period effects, such as changes in alcohol policy or availability, may also have shaped behaviors during follow-up, for example, Finland experienced many shifts in taxation and regulation in the 2000s that have influenced national consumption levels. Cohort effects may further contribute, as younger birth cohorts in Finland and other Western countries have been shown to engage in different drinking styles than older cohorts^3^. Our cohort was born from early 1940s to early 1960s, being the cohort that used to drink during early adulthood more than the previous cohorts and the most recent cohorts^3^. Although controlling for age and modeling the time in relation to retirement, the interplay of these age, period, and cohort dynamics may therefore partly account for the observed stability in socioeconomic differences in alcohol use across the retirement transition.

## Conclusion

In conclusion, this study shows that overall alcohol drinking declined and changes in socioeconomic differences in drinking patterns varied by the measure used. Retirement did not markedly change drinking patterns or socioeconomic differences.

From a policy perspective, these findings highlight the importance of monitoring alcohol use in older adults, even in the absence of sharp increases with retirement or marked changes in socioeconomic differences during ageing. Statutory retirement represents a critical window for promoting healthy lifestyle changes, and tailored interventions could help prevent escalation of harmful drinking in specific subgroups such as individuals with prior heavy use.

The socioeconomic gradients should be interpreted within the alcohol harm paradox, which highlights that although individuals in higher socioeconomic positions often drink more, those in lower socioeconomic positions experience greater alcohol-related harm at comparable or even lower levels of consumption. Thus, while higher SEP groups may exhibit higher prevalence of risky drinking, they may also be partially shielded from its adverse consequences due to their greater economic, social, and health resources.

These findings underscore that socioeconomic advantage does not uniformly translate into healthier behaviors and that the health implications of alcohol-related behaviors are shaped not only by their prevalence but also by the social and material contexts in which they occur. However, for disadvantaged groups, even modest levels of alcohol consumption may exacerbate existing vulnerabilities, contributing to widening health inequalities despite similar or lower consumption compared with higher SEP groups.

These findings underscore the importance of considering both excessive consumption among higher socioeconomic groups and alcohol-related harms disproportionately affecting disadvantaged groups. While it is important to tackle the higher and slowly declining levels of alcohol consumption in higher socioeconomic groups, it is also well-known that poorly targeted preventive interventions may widen the socioeconomic differences, which should be considered when targeting interventions. Broader public health strategies - such as maintaining alcohol affordability policies, supporting health literacy, and integrating alcohol counseling into retirement planning could further mitigate long-term risks. Ensuring that prevention efforts address both socioeconomic differences and ageing-related vulnerabilities is essential for reducing alcohol-related harm in older populations.

## Data Availability

Data can be obtained through reasonable requests. The Helsinki Health Study survey data cannot be publicly released due to stringent data protection laws and regulations. Use of the data is restricted to scientific research and collaboration with the research group's partners, subject to a reasonable request and study plan. Further details regarding the availability of the survey data can be obtained by contacting the Helsinki Health Study research group at.

