## Supplementary file for "Changes in socioeconomic differences in drinking patterns before and after retirement - a 22-year follow-up study"

Supplementary table 1. Pre- and post-retirement slopes by SEP groups among participants of the Helsinki Health Study 2000–2022 (N =4889)

| | **Occupational class** | **Slope Pre-retirement (95% CI)** | **Slope Post-retirement (95% CI)** | **Period x time (p)** |
|---|---|---|---|---|
| **Weekly doses** | Professionals | –0.051 (–0.194, 0.091) | 0.030 (–0.029, 0.055) | 0.970 |
| | Semi-professionals | 0.005 (–0.008, 0.018) | –0.006 (–0.027, 0.014) | 0.544 |
| | Manual & routine workers | –0.006 (–0.017, 0.004) | 0.015 (–0.002, 0.032) | 0.076 |
| **Non-drinking** | Professionals | 0.042 (–0.055, 0.139) | 0.032 (–0.242, 0. 050) | 0.980 |
| | Semi-professionals | 0.001 (–0.018, 0.021) | 0.015 (–0.024, 0.053) | 0.454 |
| | Manual and routine workers | –0.011 (–0.028, 0.007) | 0.008 (–0.026, 0.042) | 0.644 |
| **Binge drinking** | Professionals | –0.126 (–0.222, –0.031) | 0.069 (–0.060, 0.099) | 0.912 |
| | Semi-professionals | –0.013 (–0.040, 0.015) | 0.019 (–0.002, 0.059) | 0.369 |
| | Manual and routine workers | –0.007 (–0.030, 0.015) | 0.026 (–0.007, 0.058) | 0.122 |
